# Vayu: Screen-Free Haptic Breathwork with HRV-Adaptive Control—Pilot Outcomes and System Design

**DOI:** 10.64898/2026.06.08.26355230

**Authors:** Dhruv Adhia, Deepali Raiththa, Arron Ferguson, Philippe Pasquier

**Author notes:** Equal contribution (Dhruv Adhia, Deepali Raiththa).

## Abstract

**Vayu** is a mobile breathwork system comprising an iOS companion app and Apple Watch application that delivers slow, resonant breathing using *screen-free haptic cues*, HRV-adaptive pacing, and reflective journaling grounded in Patanjali’s five states of mind. The watchOS component provides tactile phase guidance and real-time biometric sensing (heart rate, HRV), while the iOS interface supports analytics and personalized recommendations.

In a 4–6-week naturalistic pilot involving 199 adults (ages 22–65) across Canada, the United States, and India, participants engaged in daily 5–10-minute sessions guided by on-wrist haptics. Average adherence was 4.1 ± 2.3 sessions per week, with 71% of active users maintaining at least 3 sessions per week. By week four, perceived stress (PSS-10) decreased by 2.5 points, resting heart rate declined by 7.4 bpm, and HRV increased by a median of 28.6% relative to baseline, accompanied by mood improvements. No adverse events were reported. HRV metrics are derived from Apple Watch PPG-based proxies and interpreted as relative trends. These findings suggest **Vayu** is effective and well-tolerated, demonstrating strong engagement and early efficacy signals.

## 1 Introduction

Stress, anxiety, and burnout are widespread burdens on well-being and productivity. Among accessible self-regulation practices, slow resonant breathing (≈ 0.1 Hz; ∼ 6 breaths/min) has strong physiological basis: it amplifies respiratory sinus arrhythmia (RSA), strengthens baroreflex gain, and shifts autonomic balance toward parasympathetic (vagal) dominance, supporting emotion regulation and stress recovery [2, 15, 16, 19]. Heart rate variability (HRV)—the variation in time intervals between consecutive heartbeats—is a convenient, non-invasive marker of this autonomic regulation [15].

Meta-analytic and controlled studies report meaningful reductions in self-reported distress alongside increases in HRV metrics when individuals train near their personal resonance frequency [8, 11, 16, 19, 20]. Consumer wearables now enable brief, in-situ breathing sessions paired with continuous sensing. While PPG-based devices such as Apple Watch provide reliable heart rate trends and HRV proxies, best practice treats wearables’ HRV as appropriate for within-person trend analysis rather than clinical absolutes [12, 17, 21].

Despite this opportunity, many mobile breathwork apps rely on visual cues or audio instructions [5, 13], increasing cognitive load and undermining interoceptive focus [5, 13, 14]. Prior haptic systems relied on fixed metronome rhythms without HRV-adaptive control [7, 23], and none integrate reflective journaling based on yogic cognitive states.

We **hypothesize** that subtle on-wrist haptic pacing may lower visual demands and create a more immersive breathwork experience. **Vayu** addresses these gaps through: (i) *eyes-free* on-wrist haptics encoding inhale/hold/exhale phases; (ii) on-device HR/HRV logging for progress visualization; (iii) adaptive algorithms personalizing to user routines; (iv) reflective journaling using Patanjali’s five states of mind (*Kshipta*/restless, *Mudha*/dull, *Vikshipta*/distracted, *Ekagra*/one-pointed, *Nirodha*/stilled) [1, 3, 22]; and (v) closed-loop biofeedback adjusting breathing tempo using HRV-derived coherence targets.

### 1.1 Research Questions

This pilot study addressed three research questions:

#### RQ1 Effectiveness

Does screen-free, HRV-adaptive haptic breathwork reduce perceived stress, increase HRV, and improve mood in a naturalistic setting?

#### RQ2 Adherence and Engagement

Does haptic pacing with adaptive control achieve engagement rates comparable to prior breathwork interventions?

#### RQ3 Mind States and Physiology

How do participants’ self-reported mind states change over time, and how are these changes associated with HRV and mood?

For RQ1, we hypothesized participants would show decreases in PSS-10, increases in HRV and mood. For RQ2, we hypothesized adherence rates comparable to or exceeding typical mobile health interventions. For RQ3, we hypothesized that time spent in steadier states would increase and correlate with improved HRV and mood.

## 2 Methods

### 2.1 Study Design and Participants

This was a prospective, single-arm, naturalistic pilot conducted over 4–6 weeks. Adults aged 22–65 years, residing in Canada, the United States, or India, used the Vayu app on Apple Watch Series 8 (or newer) for daily sessions lasting 5–10 minutes. Exclusions included unstable cardiopulmonary disease, pregnancy, or prior adverse responses. The final analyzed sample was *N* = 199. An a priori power analysis indicated that approximately 44 participants would be sufficient to detect a medium effect size (*d* = 0.5) with *α* = 0.05 and power 1 − *β* = 0.80. We targeted enrollment exceeding 150 to account for attrition.

### 2.2 System Architecture

**Vayu** comprises an iOS companion app and dedicated watchOS application. The Apple Watch provides PPG-based HR and device HRV (SDNN/rMSSD proxy) via HealthKit. The haptic engine delivers parameterized tap patterns encoding breathing phases: densifying pulses for inhale, sparse ticks for hold, and sparsifying pulses for exhale. Distinct double-tap patterns mark phase boundaries.

The iOS app enables users to: (i) label their current mind state, (ii) access quickstart breath training, (iii) launch curated or self-guided sessions, and (iv) view progress summaries. The app includes four curated starting points—Morning Bliss, Daytime Focus, Evening Calm, and Nighttime Relax—automatically selecting suitable breathing protocols (Box Breathing 4-4-4-4, 4-7-8 Breathing, Anulom Vilom, Bhastrika, Kapalbhati) [9, 10, 18]. The system uses a rule-based adaptive control framework with a closed-loop proportional-integral (PI) controller that adjusts breathing tempo in real time based on HRV coherence metrics.

### 2.3 Haptic Design and Implementation

The haptic engine provides eyes-free discriminability between *inhale, hold*, and *exhale* phases with low cognitive load. Each tactile pulse is a brief vibration lasting approximately 30–50 ms, with intervals of 120–180 ms or longer to prevent perceptual fusion. Two vibration strengths are used: a *strong* pulse signals phase boundaries, and a *medium* pulse guides ongoing breathing. Every transition is marked by a distinctive *double-tap*—two strong pulses, 80–120 ms apart. The inhale uses a frequency ramp from 3 Hz to 6 Hz, producing a perceptible build-up. The exhale mirrors this with a wind-down pattern. During breath-holds, low-frequency ticks (0.8–1.2 Hz) preserve timing without driving tempo.

### 2.4 Safety Constraints

We implement a two-layer safety shield that (i) prunes unsafe options before selection and (ii) monitors physiology during sessions to down-shift or stop if risk rises. A real-time monitoring system computes a hyperventilation risk index from sustained high respiratory rate, abrupt HR spikes, or user-reported dizziness/tingling. If risk exceeds thresholds, the app automatically: (a) lengthens exhale or slows tempo, (b) skips stimulating cycles, or (c) terminates with a recovery prompt. Guards cap maximum respiratory rate, consecutive stimulating cycles, hold duration, and session length, with a safe fallback always available.

### 2.5 Adaptive Control and Personalization

Tempo is guided toward each user’s resonance frequency (typically 4.5–7 breaths/min, ≈0.1 Hz) through brief paced-breathing probes. The system monitors HRV coherence—computed as the fraction of HRV power in the resonance band (0.08–0.12 Hz) relative to the broader low-to-mid frequency range (0.04–0.40 Hz)—from a sliding window (60–120 s) of interbeat intervals. When coherence falls below target thresholds, the system adjusts breathing tempo within safe bounds to help users approach their optimal resonance frequency.

For session recommendations, the system collects user context (stated goals, time of day, current mood), session history (practice frequency, technique preferences), and physiological trends (rolling HRV metrics, heart rate patterns). A hybrid recommendation engine combines rule-based matching with AI-generated suggestions to generate personalized program structures and technique sequences tailored to individual needs.

### 2.6 Outcomes and Data Collection

Primary outcomes are adherence (sessions per week) and change in perceived stress (PSS-10 scale) [6]. Secondary outcomes include resting heart rate, HRV metrics (SDNN/device-derived score), mood rating (1–5 scale), and mind state transitions. Heart rate is sampled continuously during active sessions via watchOS APIs, while HRV metrics are retrieved from HealthKit. Users log baseline and weekly measures in comparable contexts (morning, seated, ≥60 s).

We interpret HRV as a within-person metric—reflecting how an individual’s autonomic state changes relative to their own baseline. All HRV estimates are derived from the same device and firmware version. Inter-beat interval time series are processed using a standardized pipeline: (1) artifact detection and removal; (2) ectopic beat correction; (3) cubic spline interpolation; (4) resampling to 4 Hz; and (5) spectral estimation via Welch’s periodogram. We compute time-domain (SDNN, RMSSD) and frequency-domain HRV indices [12, 15]. Outliers are trimmed at 5th and 95th percentiles. For continuous endpoints, we report means±SD and pre–post changes (Week 4 vs. baseline) with effect sizes (Cohen’s *d*) and two-tailed paired *t*-tests.

## 3 Results

### 3.1 Sample, Adherence, and Quantitative Outcomes

Table 1 summarizes the cohort. Users completed on average **4.1** ± **2.3** sessions/week (median 3.8) in the active subgroup (*n* = 58); **71%** met ≥3 sessions/week. Median session duration was **8.2** ± **3.1 min**.

**Table 1:** Sample demographics and adherence.

| Characteristic | Value |
| --- | --- |
| Total $N$ | 199 |
| Age | 22–65 years (adults) |
| Geography | Canada, United States, India |
| Device | Apple Watch Series 8+ |
| Mean sessions/week (active) | $4.1 \pm 2.3$ |
| Users with $\geq 3$ sessions/week | 71% |
| Median session length | $8.2 \pm 3.1$ min |

Table 2 summarizes main endpoints from baseline to week 4, showing a median HRV improvement of 28.6%, a 10.4% decline in resting HR, a 2.5-point decrease in PSS-10 (15%), and a 0.4-point increase in mood rating.

**Table 2:** Main outcomes from baseline to Week 4 (intent-to-treat, available cases).

| Outcome | $n$ | Baseline | Week 4 | $\Delta$ (95% CI) | Cohen’s $d$ |
| --- | --- | --- | --- | --- | --- |
| PSS-10 (0–40) | 199 | $16.2 \pm 6.1$ | $13.7 \pm 5.7$ | $-2.5 [-3.33, -1.67]$ | 0.41 |
| HRV (SDNN/score) | 199 | $37.8 \pm 12.4$ | $54.5 \pm 17.2$ | $+16.7 [14.55, 18.85]$ | 0.68 |
| Resting HR (bpm) | 199 | $71.3 \pm 8.5$ | $63.9 \pm 7.9$ | $-7.4 [-8.55, -6.25]$ | 0.86 |
| Mood rating (1–5) | 199 | $2.8 \pm 0.9$ | $3.2 \pm 1.0$ | $+0.4 [0.27, 0.53]$ | 0.34 |

*Notes.* Two-tailed paired $t$ -tests; $p < 0.001$ for all outcomes. HRV percentage change reported as median (28.6%). All $p$ -values are descriptive given single-arm design.

### 3.2 State Transitions and Associations

Session-end user reports assigned state labels based on Patanjali’s framework. We analyzed transitions using a Markov transition matrix *P*_*ij*_ summarizing probabilities of moving from one state to another (Figure 4). At the group level, dwell in steadier states (Vikshipta and Ekagra) increased over time, while the proportion in more scattered states (Kshipta and Mudha) decreased. The diagonal of the transition matrix shows how often participants remain in the same state across sessions: Kshipta 29%, Mudha 28%, Vikshipta 36%, Ekagra 51%, and Nirodha 64%. The probability of progressing to a more focused state is 43% from Kshipta, 44% from Mudha, 38% from Vikshipta, and 29% from Ekagra.

**Figure 1:**
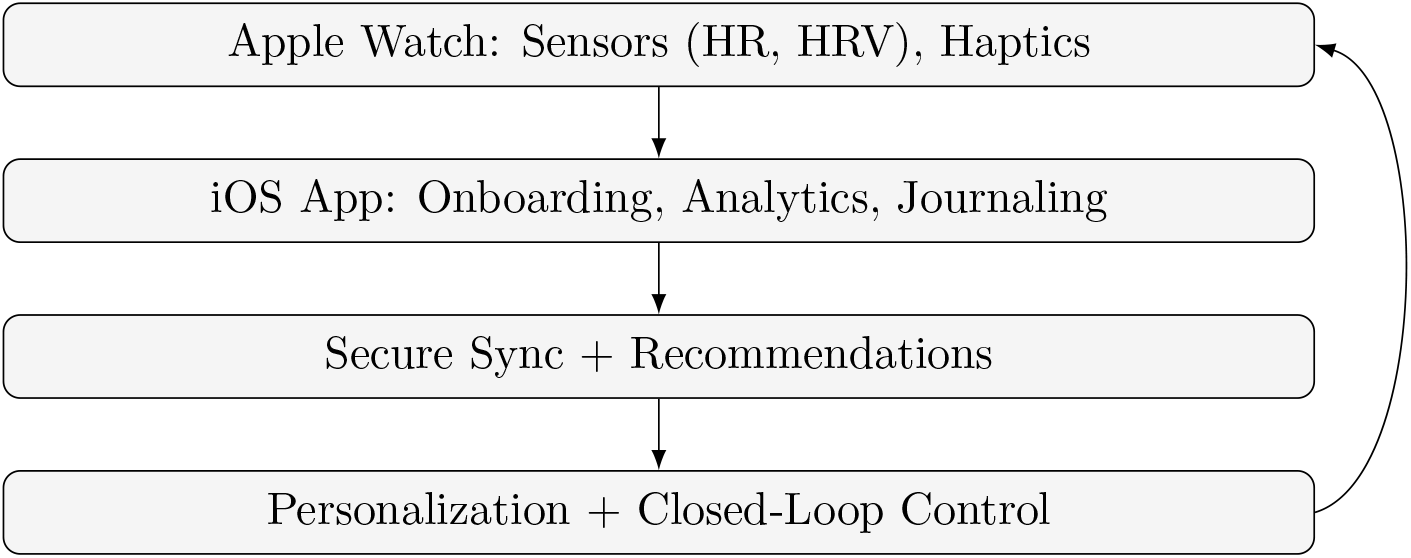
End-to-end pipeline: sensing, analytics, personalization, and on-wrist guidance.

**Figure 2:**
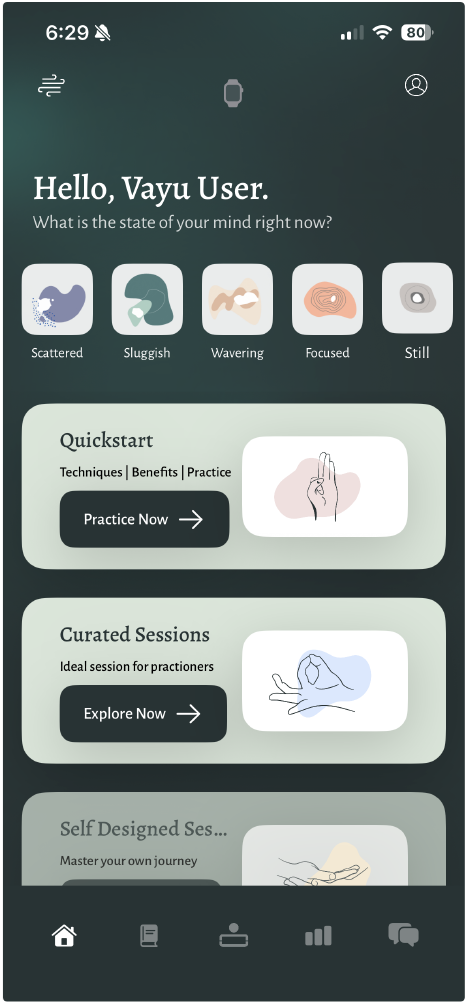
Vayu home screen.

**Figure 3:**
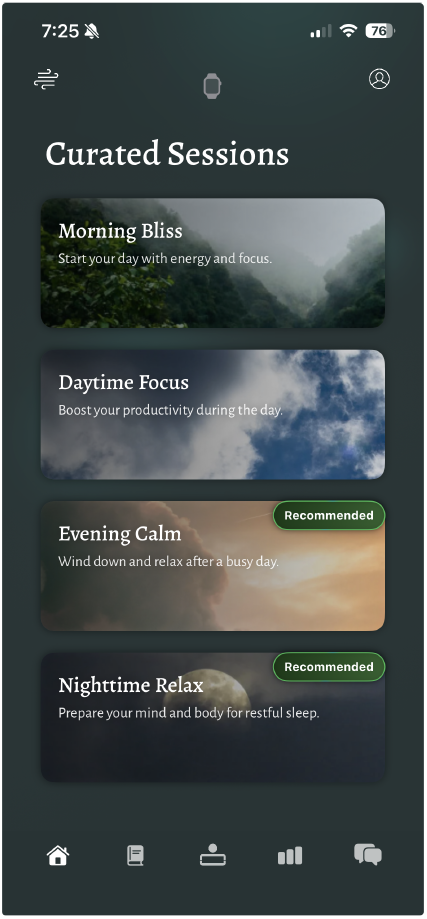
Curated sessions UI.

**Figure 4:**
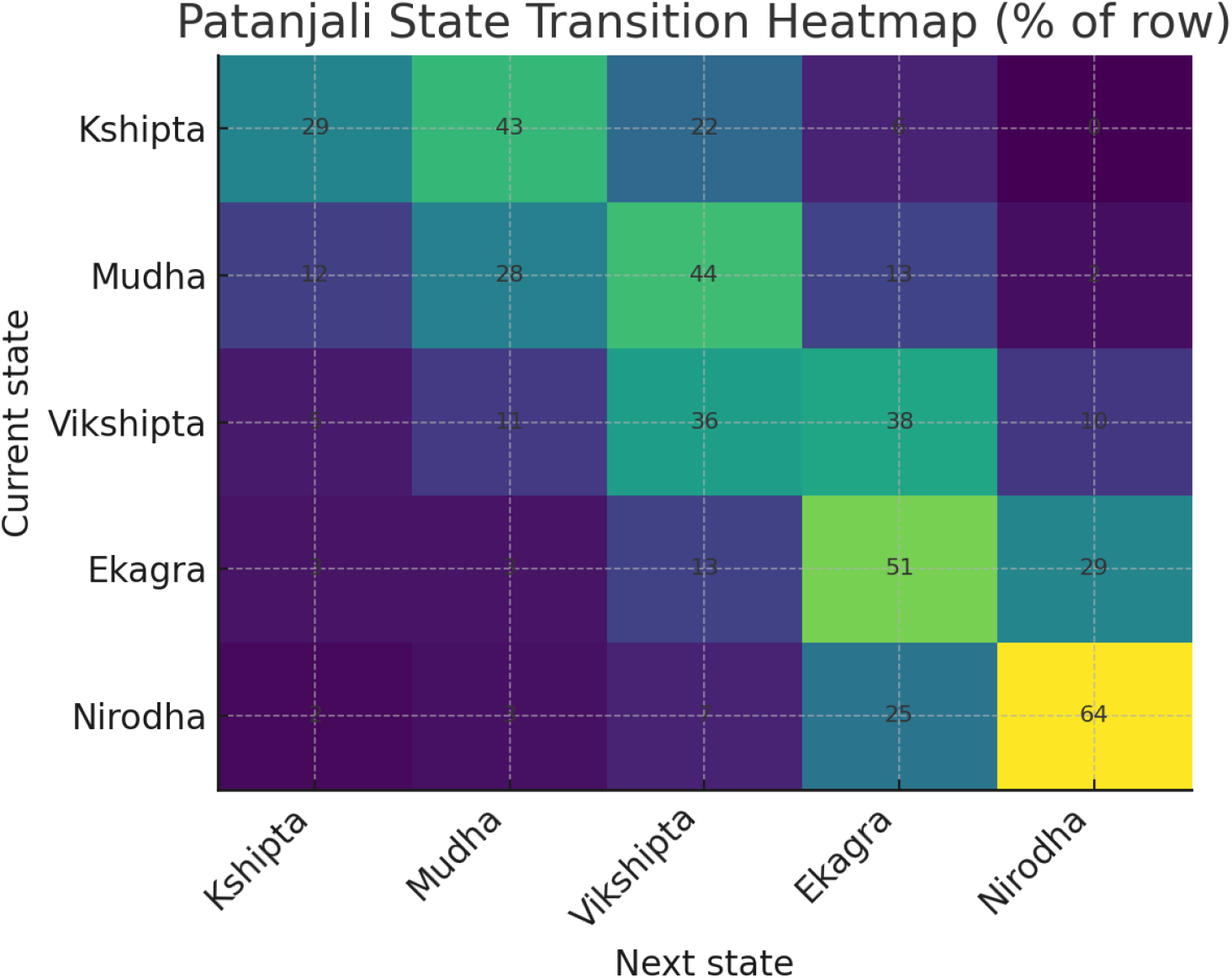
Patanjali state-transition heatmap. Each row is the current state; cell values show the percentage of next-state transitions (row-normalized).

Across sessions, higher HRV co-occurred with steadier states [2], and week-over-week increases in dwell time for Vikshipta and Ekagra were associated with HRV gains. Participants meeting the adherence target (≥ 3 sessions/week) showed larger mood improvements. All associations are observational and do not imply causality.

### 3.3 Qualitative Feedback

Participants reported immediate post-session increases in calmness, measured on a 0–10 scale. Many highlighted the value of eyes-free haptic guidance, noting it reduced cognitive load compared to visual timers. Short session formats (5–10 minutes) were appreciated for fitting into daily routines. Visible feedback on HR/HRV progress was cited as motivating.

## 4 Discussion

In a naturalistic, multi-country cohort, Vayu achieved adherence of ∼4 sessions/week with measurable improvements in PSS-10, HRV, HR, and mood within 4 weeks. These magnitudes align with instructor-led HRV-biofeedback literature [4, 8, 11, 20], despite entirely at-home delivery. The median 28.6% increase in HRV and 7.4 bpm reduction in resting heart rate suggest meaningful autonomic improvements (increased vagal activity, reduced sympathetic load). The 2.5-point reduction in PSS-10 represents approximately 15% improvement relative to baseline (*d* = 0.41). Mood improvements were modest but consistent (*d* = 0.34).

Journaling transitions suggest cognitive-affective alignment with training: participants increasingly reported steadier mind states over time, with these states co-occurring with higher HRV. The combination of haptics (low cognitive load), feedback, and reflective practice likely contributed to engagement and outcomes. The 71% adherence rate (achieving ≥3 sessions/week) compares favorably to typical mobile health app engagement rates [13].

### 4.1 Limitations

The single-arm design and potential self-selection introduce bias and limit causal inference. Consumer wearables estimate HRV, which may not match gold-standard ECG validity. Apple Watch HR and HRV trends are suitable for within-person *relative* change but can underestimate absolute HRV. Adherence data were self-reported. Metadata on participant sex and device model were incomplete. Naturalistic confounders (sleep, caffeine, physical activity) could not be fully controlled. The single-arm design cannot rule out expectancy effects, regression to the mean, or Hawthorne effects. Without a control group, we cannot determine whether improvements stem from the haptic guidance, the breathing practice itself, increased self-awareness, or mere participation. Causality requires randomized controlled trials with active controls, pre-registration, and blinded analyses.

### 4.2 Implications and Future Directions

These findings suggest that eyes-free haptic breathwork delivered via consumer wearables is feasible and well-tolerated, with promising adherence and early efficacy signals. The haptics-first approach may reduce cognitive load compared to screen-based guidance, supporting interoceptive focus. The integration of Patanjali’s framework provides a structured phenomenological scaffold for self-awareness that aligns with the physiological training. The absence of adverse events is encouraging given concerns about unsupervised breath training. The built-in safety guards help mitigate risks in unsupervised digital health interventions.

Building on pilot findings, we plan a prospective, 8–12 week randomized controlled trial comparing the full Vayu system with an active control (timer-paced breathing without haptics). Primary outcome will be PSS-10 change, with secondary outcomes including HRV, resting HR, sleep quality (PSQI), and well-being (WHO-5). Future work should evaluate clinical and psychophysiological benefits in controlled settings, particularly for populations with elevated baseline stress or anxiety.

## 5 Conclusion

A haptics-centric, feedback-enabled breathwork mobile application demonstrated robust user adherence in naturalistic settings, yielding significant improvements in stress reduction, HRV, heart rate, and mood over 4 weeks. The eyes-free haptic approach shows promise for accessible, engaging digital breathwork. This work offers methodological, theoretical, and design contributions. *Methodologically*, it demonstrates how to integrate real-time physiological adaptation into a naturalistic pilot and illustrates best practices for preprocessing wearable HRV data. *Theoretically*, it bridges yogic cognitive-state frameworks with measurable psychophysiological markers, providing a novel lens for understanding how subjective experience relates to autonomic regulation. *Design-wise*, the project offers replicable guidelines for developing eyes-free biofeedback systems that use haptics and adaptive pacing to support self-regulation in everyday contexts. Progression to randomized controlled trials will be essential to rigorously evaluate therapeutic potential and establish efficacy as a digital health intervention.

## Declaration of Conflicting Interests

Deepali Raithatha is affiliated with Prana Labs Inc., the company that developed the Vayu application. The other authors declare no potential conflicts of interest with respect to the research, authorship, and/or publication of this article.

## Funding

The authors received no financial support for the research, authorship, and/or publication of this article.

## Ethics Statement

Ethical approval was granted by the Research Ethics Board at Simon Fraser University (SFU REB approval #30003493). All participants provided informed consent at onboarding.

## Guarantor

Dhruv Adhia.

## Contributorship

DA and DR conceived and designed the study. DR developed the Vayu application. DA conducted data analysis. AF contributed to methodology and manuscript preparation. PP provided supervision and critical revision. All authors reviewed and approved the final manuscript.

## Data Availability

The datasets generated and analyzed during the current study are not publicly available due to privacy restrictions and proprietary considerations but are available from the corresponding author on reasonable request for academic research purposes, subject to appropriate data use agreements.

## Acknowledgment

The authors acknowledge the use of Perplexity AI (Deep Research mode) for literature identification and Grammarly for language refinement. The authors reviewed all output and take full responsibility for content.

## Notes

### Competing Interest Statement

Deepali Raithatha and Dhruv Adhia is affiliated with Prana Labs Inc., the company that developed the Vayu application. The other authors declare no potential conflicts of interest with respect to the research, authorship, and/or publication of this article.

### Author Declarations

Research Ethics Board of Simon Fraser University gave ethical approval for this work.

### Summary of Updates

Corrected name of the authors to Arron instead of Aaron.

